# Decisional Needs and Patient Treatment Preferences for Heart Failure Medications: Scoping Review Protocol

**DOI:** 10.1101/2022.07.05.22277267

**Authors:** Ricky D. Turgeon, Arden R. Barry, Blair MacDonald

## Abstract

**Introduction:** Treatment decisions regarding heart failure with reduced ejection fraction (HFrEF; ejection fraction ≤40%) pharmacotherapy are complex. Decision aids can bridge this knowledge-to-practice gap and improve the integration of patients’ preferences and values for patient-centered care. However, little is known about the preferences and decisional needs of patients regarding these medications. The objectives of this scoping review are to identify, map and synthesize the literature evaluating the decisional needs, treatment preferences and values of patients making decisions regarding HFrEF medications.

**Methods and Analysis:** We will search MEDLINE, Embase, CINAHL (inception-April 2022), bibliographies of included studies and relevant reviews, Web of Science ‘cited references’, cocites.com, clinicaltrials.gov, Epistemonikos, and the Ottawa Decision Aid Inventory, without language restriction. We will include qualitative, quantitative, and mixed-methods studies that describe patient and clinician decisional needs, or patient treatment preferences or values regarding HFrEF medications guided by the Ottawa Decision Support Framework, or decision aids to support HFrEF medication decisions. One author will perform all searches and upload results to Covidence. Two review authors will independently screen retrieved article titles and abstracts for inclusion, review full-text for final inclusion, and extract data from included articles and decision aids using a standardized data extraction form. We will present a graphical abstract mapping what is known about decisional needs and patient preferences and values, decisional support interventions, and decisional outcomes regarding HFrEF medications. We will also describe extracted data in narrative and tabular format to address the scoping review objectives, and discuss implications for practice and subsequent research in the field of shared decision-making for HFrEF.

**Ethics and Dissemination:** Research ethics board approval is not required for this scoping review of published data. We will present the findings at relevant conferences, publish a peer-reviewed manuscript, and disseminate results via institutional and partner social media platforms.

**Strengths and limitations of this study:**

- This scoping review will systematically identify, map and synthesize the decisional needs, treatment preferences and values of patients regarding heart failure medication decisions.
- We have developed a comprehensive search strategy to identify qualitative, quantitative and mixed-methods studies (published and unpublished), as well as decision aids.
- The review will follow methodology outlined in the Joanna Briggs Institute Manual for Evidence Synthesis and the Preferred Reporting Items for Systematic Reviews and Meta-Analyses Extension for Scoping Reviews reporting guidelines.
- The results of the scoping review will ultimately be limited by available studies; however, preliminary searches have identified several eligible studies.

## Introduction

### The complexity of pharmacotherapy for heart failure

Heart failure (HF) affects an estimated 64.3 million people globally, impairs quality of life, and has a lower survival than most common cancers.[1–4] HF is primarily classified based on left ventricular ejection fraction, or the percentage of blood inside the left ventricle that is pumped out with each contraction, categorized as preserved (ejection fraction ≥50%), mildly reduced (41-49%), or reduced (≤40%).[3,5] Most HF medications target HF with reduced ejection fraction (HFrEF), which accounts for approximately half of all HF cases.[2–4,6]

The current guideline-directed management strategy for HFrEF pharmacotherapy focuses on the routine combination of four core medications, followed by titration of these medications and subsequent consideration of up to six additional medications based on patient-specific clinical factors.[4,7] The use of guideline-directed HFrEF medications is low in clinical practice despite high-quality evidence that they improve survival and quality of life.[7–12] In a contemporary cohort from the United States, only 22% of HFrEF patients were simultaneously prescribed all guideline-recommended medications.[8] Similar treatment gaps have been identified globally.[13,14] Even when these medications are prescribed, they are seldom titrated to evidence-based doses,[8–10] and patient non-adherence is common.[11,12] Knowledge gaps, including inaccurate perceptions of pharmacotherapy benefits and harms, are commonly-reported barriers to HF care.[15]

### Shared decision-making (SDM) and HFrEF pharmacotherapy

Decision aids can bridge this knowledge-to-practice gap and improve the integration of patients’ preferences and values for patient-centered care. The SDM model of care emphasizes the joint participation of patients and clinicians in making healthcare decisions.[16] In SDM, the clinician’s role is to provide accurate and understandable information about available options for a given decision, elicit patient preferences, and help patients choose the option that best fits with their values and preferences.[16] Decision aids are educational tools that facilitate SDM by providing clear evidence-based information about treatment options, help patients elicit their values and preferences, and aid in the deliberation of healthcare decisions among available options.[17] Numerous studies across health decisions have demonstrated that the use of decision aids improves patient knowledge about their risks and options, increases readiness to make a decision, helps them choose an option reflecting their preferences and values, and increases satisfaction with their care.[17]

The abundance of available HFrEF pharmacotherapy options has resulted in a renewed call for novel strategies to facilitate SDM for HFrEF pharmacotherapy, including interactive decision aids that provide individualized estimates of benefits and harms.[18–20] Most recently, the 2021 *American College of Cardiology Consensus Decision Pathway for Optimization of Heart Failure Treatment* advocates for SDM and use of decision aids for HFrEF pharmacotherapy decisions when available.[7] To date, integration of SDM has been limited to decisions regarding device therapy and palliative care in patients with advanced HF.[21–23] We are aware of only one decision aid addressing a single HFrEF medication class,[24] and there is no broadly-available decision aid that incorporates contemporary HFrEF medication options.

### Theoretical framework to support the development of a decision aid

Decision aids based on the validated and widely-used Ottawa Decision Support Framework (ODSF) can address decisional needs, improve decision quality, and lead to better health outcomes.[17,25–27] The ODSF posits that “decisional support interventions (such as decision aids) that address decisional needs may increase the quality of the decision,” along with “positive downstream effects on actions […] as well as achievement of values-based health outcomes,”[27] as supported by empirical evidence for decision aids.[17] A recent systematic review by Hoefel et al. identified the following common decisional needs of people making health decisions: decisional conflict, inadequate knowledge, unrealistic expectations, unclear values, inadequate support and resources, complex decision characteristics, and individual needs stemming from patient factors such as age, gender, ethnicity, or other social determinants of health.[25] Although common, not all of the above decisional needs manifest for every health decision, and it remains unclear which of these decisional needs are important in decisions regarding HFrEF pharmacotherapy.

Important recent work has been done to understand patient preference regarding HFrEF medications; however, these are insufficient to inform the development of a decision aid. Studies evaluating treatment preferences of HFrEF patients have focused almost exclusively on the trade-off between maximizing survival versus quality of life, where patients have strong preferences to maximize one or the other.[28–31] However, many HFrEF medications improve both survival and quality of life,[4,32] and trade-offs may come in the form of other considerations such as side-effects, cost, and pill burden.[33–35] Additionally, studies may provide data that could inform decisional needs according to the 2020 ODSF, but not code them as such due to use of an alternate theoretical framework to inform their analyses.[36,37]

### Rationale for a scoping review

Decision aids must be informed by the decisional needs, treatment preferences and values of patients and clinicians, which are largely unknown for decisions regarding HFrEF pharmacotherapy. A preliminary search of PubMed, MEDLINE, the Cumulative Index to Nursing and Allied Health Literature (CINAHL), and JBI Evidence Synthesis (inception to August 24, 2021) identified a mix of qualitative and quantitative studies exploring decisional needs and treatment preferences for medical and invasive therapies in patients with HFrEF, and did not identify any scoping or systematic reviews (published or underway) on this topic. Given the anticipated scarcity and heterogeneity of information on this topic, a scoping review method was chosen in order to systematically assess and synthesize knowledge on decisional needs of HFrEF patients and their clinicians when making decisions about HFrEF medications, and patient preferences and values regarding HFrEF medications.

### Objectives

The main objectives of this scoping review are to identify, map and synthesize the literature evaluating the decisional needs, treatment preferences and values of HFrEF patients making treatment decisions.

### Review questions

Specifically, we aim to address the following questions:

1. What methodological designs have been used to assess HFrEF patient and clinician decisional needs, preferences and values regarding HFrEF medication decisions?
2. What decisional needs have been identified as relevant to HFrEF patients and their clinicians when making decisions regarding HFrEF medications?
3. What treatment attributes do patients and clinicians consider (or wish to consider) when making decisions regarding HFrEF medications?
4. What are patients’ treatment preferences and values regarding treatment attributes relevant to HFrEF medications?
5. What are the key characteristics of existing patient decision aids for HFrEF medications?

The findings from this scoping review will inform the need for and design of a comprehensive HFrEF medication decisional needs assessment, and subsequent development of a patient decision aid.

## Methods and Analysis

We will conduct this scoping review based on methodology outlined in the Joanna Briggs Institute Manual for Evidence Synthesis, and report the findings according to the Preferred Reporting Items for Systematic Reviews and Meta-Analyses Extension for Scoping Reviews (PRISMA-ScR).[38,39]

### Eligibility criteria

#### Participants

We will primarily consider studies including patients with HFrEF, with “reduced ejection fraction” defined as an ejection fraction of 40% or less, as well as clinicians.[5] To account for changes in the definition of “reduced ejection fraction” over time, we will also include studies using alternate thresholds (e.g. <50%). We will exclude studies limited to patients with advanced HF being considered for mechanical circulatory support or heart transplant, as well as studies focusing on palliative care, end-of-life care, or advanced-care planning.

#### Concept and Context

The concept of interest for this scoping review will be decisional needs, treatment preferences, and values regarding HFrEF medications, as well as decisional aids developed to support HFrEF medication decisions. We will also consider studies assessing decisional needs and preferences regarding other therapies for HFrEF, including medical devices, interventional procedures, and surgeries other than mechanical circulatory support or heart transplant, if the needs/preferences may apply to HFrEF medication decisions (e.g. assess preferences around quality of life versus cost; assess decisional needs around information deficiency/overload).

#### Design

We will consider qualitative (e.g. interviews, focus groups), quantitative (e.g. surveys), and mixed-methods studies available as full-text articles or conferences abstract, as well as decision aids. We will not restrict eligibility by language (articles not in English or French will be translated using Google Translate or, if required, a formal translation service). We will exclude reviews, editorials, commentaries, protocols/design papers, and non-research letters.

### Information sources and search

The search strategy will aim to locate both published and unpublished studies and decision aids. We will conduct detailed literature searches of the following databases from inception to April 1, 2022: MEDLINE, Embase, and CINAHL. We performed an initial limited search of MEDLINE to identify articles on the topic, and used words contained in the titles and abstracts of relevant articles, along with the index terms used to describe the articles, to develop a full search strategy. Online supplemental appendix 1 outlines the full electronic search strategy for all databases. Sources of unpublished or grey literature will include manual search of bibliographies of included studies and relevant reviews, and searches of Web of Science ‘cited references’, cocites.com, clinicaltrials.gov, Epistemonikos, and the Ottawa Decision Aid Inventory (decisionaid.ohri.ca/AZinvent.php).

### Selection of sources of evidence

We will upload search results to Covidence (Veritas Health Innovation, Melbourne, Australia) and remove duplicates. Following a pilot test by both reviewers to ensure consistency in interpretation and application of the eligibility criteria, two review authors will independently screen retrieved article titles and abstracts for inclusion, followed by full-text review for final inclusion. Disagreements will be resolved by discussion and consensus. We will document study selection in a PRISMA-ScR flow diagram, with reasons for exclusion for articles excluded during the full-text review.

### Data charting process and data items

Two authors will independently extract data from included articles and decision aids using a standardized data extraction form developed by the reviewers (online supplemental appendix 2 for draft study data extraction form). We will resolve any disagreements that arise through discussion, and by consulting a third reviewer if needed. We will modify the data extraction tool as required during data extraction and describe revisions in detail in the full scoping review report. In cases where pertinent information is not reported, we will contact study authors for additional information. For decision aids, we will extract data on the developer/funder, year last updated, format, decision being made, options considered, and elements required to evaluate each decision aid using the International Patient Decision Aid Standards instrument (IPDASi) checklist.[40]

### Synthesis plan

#### Data presentation

We will present a graphical abstract mapping what is known about decisional needs and patient preferences and values, decisional support interventions, and decisional outcomes regarding HFrEF medications based on the Ottawa Decision Support Framework and other decisional frameworks used in included studies.[27] We will present extracted data narratively and in tabular form in a format that addresses the posed research questions, and discuss implications for practice and subsequent research in the field of SDM for HFrEF. Specifically, we will graphically map:

- Decisional needs according to the ODSF classification,[27] identifying which are relevant to HFrEF medication decision-making, irrelevant, or unexplored in existing studies;
- Treatment attributes that have been identified by patients and clinicians as relevant to decisions regarding HFrEF medications, along with preference weights, rating or ranking available from existing studies, and identify attributes that have not been explored in existing studies.

#### Patient and public involvement

Two patient partners were formally involved in defining and prioritizing the research questions.

## Data Availability

All data produced in the present work are contained in the manuscript.

## Ethics and Dissemination

Research ethics board approval for the conduct of this scoping review is not required as this study will involve secondary analysis of publicly-available, de-identified data. We will present the findings at relevant conferences, publish a manuscript in a peer-reviewed journal, and further disseminate results via institutional and partner social media and websites.

## Authors’ contributions

RDT conceived and wrote the first draft of the protocol. ARB contributed to the design of the protocol. ARB and BM provided critical review of the protocol content.

## References

1 GBD 2017 Disease and Injury Incidence and Prevalence Collaborators. Global, regional, and national incidence, prevalence, and years lived with disability for 354 diseases and injuries for 195 countries and territories, 1990-2017: a systematic analysis for the Global Burden of Disease Study 2017. Lancet 2018;392:1789–858. doi:10.1016/S0140-6736(18)32279-7

2 Yancy CW, Jessup M, Bozkurt B, et al. 2013 ACCF/AHA guideline for the management of heart failure: executive summary: a report of the American College of Cardiology Foundation/American Heart Association Task Force on practice guidelines. Circulation 2013;128:1810–52. doi:10.1161/CIR.0b013e31829e8807

3 Ezekowitz JA, O’Meara E, McDonald MA, et al. 2017 Comprehensive Update of the Canadian Cardiovascular Society Guidelines for the Management of Heart Failure. Canadian Journal of Cardiology 2017;33:1342–433. doi:10.1016/j.cjca.2017.08.022

4 McDonald M, Virani S, Chan M, et al. CCS/CHFS Heart Failure Guidelines Update: Defining a New Pharmacologic Standard of Care for Heart Failure With Reduced Ejection Fraction. Canadian Journal of Cardiology 2021;37:531–46. doi:10.1016/j.cjca.2021.01.017

5 Bozkurt B, Coats AJ, Tsutsui H, et al. Universal Definition and Classification of Heart Failure: A Report of the Heart Failure Society of America, Heart Failure Association of the European Society of Cardiology, Japanese Heart Failure Society and Writing Committee of the Universal Definition of Heart Failure. J Card Fail 2021;:S1071-9164(21)00050-6. doi:10.1016/j.cardfail.2021.01.022

6 Yancy CW, Jessup M, Bozkurt B, et al. 2017 ACC/AHA/HFSA Focused Update of the 2013 ACCF/AHA Guideline for the Management of Heart Failure: A Report of the American College of Cardiology/American Heart Association Task Force on Clinical Practice Guidelines and the Heart Failure Society of America. J Am Coll Cardiol 2017;70:776–803. doi:10.1016/j.jacc.2017.04.025

7 Writing Committee, Maddox TM, Januzzi JL, et al. 2021 Update to the 2017 ACC Expert Consensus Decision Pathway for Optimization of Heart Failure Treatment: Answers to 10 Pivotal Issues About Heart Failure With Reduced Ejection Fraction: A Report of the American College of Cardiology Solution Set Oversight Committee. J Am Coll Cardiol 2021;77:772–810. doi:10.1016/j.jacc.2020.11.022

8 Greene SJ, Butler J, Albert NM, et al. Medical Therapy for Heart Failure With Reduced Ejection Fraction: The CHAMP-HF Registry. J Am Coll Cardiol 2018;72:351–66. doi:10.1016/j.jacc.2018.04.070

9 Greene SJ, Fonarow GC, DeVore AD, et al. Titration of Medical Therapy for Heart Failure With Reduced Ejection Fraction. J Am Coll Cardiol 2019;73:2365–83. doi:10.1016/j.jacc.2019.02.015

10 Fiuzat M, Ezekowitz J, Alemayehu W, et al. Assessment of Limitations to Optimization of Guideline-Directed Medical Therapy in Heart Failure From the GUIDE-IT Trial: A Secondary Analysis of a Randomized Clinical Trial. JAMA Cardiol 2020;5:757–64. doi:10.1001/jamacardio.2020.0640

11 George J, Shalansky SJ. Predictors of refill non-adherence in patients with heart failure. Br J Clin Pharmacol 2007;63:488–93. doi:10.1111/j.1365-2125.2006.02800.x

12 Wu J-R, Moser DK, Lennie TA, et al. Medication adherence in patients who have heart failure: a review of the literature. Nurs Clin North Am 2008;43:133–53; vii–viii. doi:10.1016/j.cnur.2007.10.006

13 Komajda M, Anker SD, Cowie MR, et al. Physicians’ adherence to guideline-recommended medications in heart failure with reduced ejection fraction: data from the QUALIFY global survey. Eur J Heart Fail 2016;18:514–22. doi:10.1002/ejhf.510

14 Fuery MA, Chouairi F, Januzzi JL, et al. Intercountry Differences in Guideline-Directed Medical Therapy and Outcomes Among Patients With Heart Failure. JACC Heart Fail 2021;9:497–505. doi:10.1016/j.jchf.2021.02.011

15 McEntee ML, Cuomo LR, Dennison CR. Patient-, provider-, and system-level barriers to heart failure care. J Cardiovasc Nurs 2009;24:290–8. doi:10.1097/JCN.0b013e3181a660a0

16 Ting HH, Brito JP, Montori VM. Shared Decision Making: Science and Action. Circ Cardiovasc Qual Outcomes 2014;7:323–7. doi:10.1161/CIRCOUTCOMES.113.000288

17 Stacey D, Légaré F, Lewis K, et al. Decision aids for people facing health treatment or screening decisions. Cochrane Database Syst Rev 2017;4:CD001431. doi:10.1002/14651858.CD001431.pub5

18 Goyal P. Do We Need Shared Decision Making When Prescribing Guideline-Directed Medical Therapy? J Card Fail 2019;25:701–2. doi:10.1016/j.cardfail.2019.08.016

19 Enard KR, Hauptman PJ. Heart Failure, Shared Decision-making, and Social Determinants of Health: An Upstream Perspective. JAMA Cardiol 2019;4:609–10. doi:10.1001/jamacardio.2019.1763

20 Bhatt AS, Choudhry NK. Evidence-Based Prescribing and Polypharmacy for Patients With Heart Failure. Ann Intern Med 2021;174:1165–6. doi:10.7326/M21-1427

21 Allen LA, McIlvennan CK, Thompson JS, et al. Effectiveness of an Intervention Supporting Shared Decision Making for Destination Therapy Left Ventricular Assist Device: The DECIDE-LVAD Randomized Clinical Trial. JAMA Intern Med 2018;178:520–9. doi:10.1001/jamainternmed.2017.8713

22 Allen LA, Stevenson LW, Grady KL, et al. Decision making in advanced heart failure: a scientific statement from the American Heart Association. Circulation 2012;125:1928–52. doi:10.1161/CIR.0b013e31824f2173

23 Kane PM, Murtagh FEM, Ryan K, et al. The gap between policy and practice: a systematic review of patient-centred care interventions in chronic heart failure. Heart Fail Rev 2015;20:673–87. doi:10.1007/s10741-015-9508-5

24 Finnegan-Fox G, Allen LA, Jenkins A, et al. A Decision Aid for Renin-Angiotensin Inhibitor Drug Options for Patients with Heart Failure. 2017.https://www.cardiosmart.org/docs/default-source/assets/decision-aid/heart-failure-drug-options.pdf?sfvrsn=aaff9c98_1

25 Hoefel L, O’Connor AM, Lewis KB, et al. 20th Anniversary Update of the Ottawa Decision Support Framework Part 1: A Systematic Review of the Decisional Needs of People Making Health or Social Decisions. Med Decis Making 2020;40:555–81. doi:10.1177/0272989X20936209

26 Ottawa Decision Support Framework (ODSF). Ottawa Hospital Ressearch Institute. https://decisionaid.ohri.ca/odsf.html (accessed 1 Dec 2020).

27 Stacey D, Légaré F, Boland L, et al. 20th Anniversary Ottawa Decision Support Framework: Part 3 Overview of Systematic Reviews and Updated Framework. Med Decis Making 2020;40:379–98. doi:10.1177/0272989X20911870

28 Lewis EF, Johnson PA, Johnson W, et al. Preferences for quality of life or survival expressed by patients with heart failure. J Heart Lung Transplant 2001;20:1016–24.

29 Stevenson LW, Hellkamp AS, Leier CV, et al. Changing preferences for survival after hospitalization with advanced heart failure. J Am Coll Cardiol 2008;52:1702–8. doi:10.1016/j.jacc.2008.08.028

30 Havranek EP, McGovern KM, Weinberger J, et al. Patient preferences for heart failure treatment: utilities are valid measures of health-related quality of life in heart failure. J Card Fail 1999;5:85–91.

31 Kraai IH, Vermeulen KM, Luttik MLA, et al. Preferences of heart failure patients in daily clinical practice: quality of life or longevity? Eur J Heart Fail 2013;15:1113–21. doi:10.1093/eurjhf/hft071

32 Turgeon RD, Barry AR, Hawkins NM, et al. Pharmacotherapy for Heart Failure with Reduced Ejection Fraction and Health-Related Quality of Life: Systematic Review and Meta-Analysis. Eur J Heart Fail Published Online First: 25 February 2021. doi:10.1002/ejhf.2141

33 Pisa G, Eichmann F, Hupfer S. Assessing patient preferences in heart failure using conjoint methodology. PPA 2015;:1233. doi:10.2147/PPA.S88167

34 Hauber AB, Obi EN, Price MA, et al. Quantifying the relative importance to patients of avoiding symptoms and outcomes of heart failure. Current Medical Research and Opinion 2017;33:2027–38. doi:10.1080/03007995.2017.1355782

35 Heen AF, Vandvik PO, Brandt L, et al. A framework for practical issues was developed to inform shared decision-making tools and clinical guidelines. J Clin Epidemiol 2021;129:104–13. doi:10.1016/j.jclinepi.2020.10.002

36 Samsky MD, Lin L, Greene SJ, et al. Patient Perceptions and Familiarity With Medical Therapy for Heart Failure. JAMA Cardiol 2020;5:292–9. doi:10.1001/jamacardio.2019.4987

37 Smith GH, Shore S, Allen LA, et al. Discussing Out-of-Pocket Costs With Patients: Shared Decision Making for Sacubitril-Valsartan in Heart Failure. J Am Heart Assoc 2019;8:e010635. doi:10.1161/JAHA.118.010635

38 Peters M, Godfrey C, McInerney P, et al. Scoping Reviews (2020 version). In: JBI Manual for Evidence Synthesis. JBI 2020. https://synthesismanual.jbi.global (accessed 18 Aug 2021).

39 Tricco AC, Lillie E, Zarin W, et al. PRISMA Extension for Scoping Reviews (PRISMA-ScR): Checklist and Explanation. Ann Intern Med 2018;169:467–73. doi:10.7326/M18-0850

40 Elwyn G, O’Connor AM, Bennett C, et al. Assessing the quality of decision support technologies using the International Patient Decision Aid Standards instrument (IPDASi). PLoS One 2009;4:e4705. doi:10.1371/journal.pone.0004705

