## Supplement for "Decisional Needs and Patient Treatment Preferences for Heart Failure Medications: Scoping Review Protocol"

### Supplemental Appendix 1. Database search queries (database inception to April 1, 2022)

| MEDLINE (Ovid) | Embase (Ovid) | CINAHL (EBSCOhost) |
|---|---|---|
| 1. Decision Making/<br>2. Shared Decision Making/<br>3. shared decision making.ti,ab.<br>4. Patient Preference/<br>5. Patient Participation/<br>6. ((patient* or individuals) adj3 (choice* or choose* or choosing or decision* or deliberation* or dialog* or engag* or expectation* or input or perspective* or preference* or prioriti?ation or prioriti?e* or prioriti?ing or value* or voice)).ti.<br>7. (decision* or decided or decides or deciding or choice*).ti.<br>8. (decision* need*).ti,ab,kf.<br>9. Ottawa decision support framework.mp.<br>10. Clinical Decision Support System/<br>11. Decision Support Techniques/<br>12. decision aid.ti,ab.<br>**13. or/1-12** | 1. Decision Making/<br>2. Decision Making, Shared/<br>3. shared decision making.ti,ab.<br>4. Patient Preference/<br>5. Patient Participation/<br>6. ((patient* or individuals) adj3 (choice* or choose* or choosing or decision* or deliberation* or dialog* or engag* or expectation* or input or perspective* or preference* or prioriti?ation or prioriti?e* or prioriti?ing or value* or voice)).ti.<br>7. (decision* or decided or decides or deciding or choice*).ti.<br>8. (decision* need*).ti,ab.<br>9. Ottawa decision support framework.mp.<br>10. Decision Support Systems, Clinical/<br>11. Decision Support Techniques/<br>12. decision adj3 (support* OR aid* OR navigation OR patient* OR tool*).ti,ab.<br>**13. or/1-12** | 1. ((MH "Patient Preference") OR patient preference* OR patient priorit* OR (MH "Values Clarification") OR patient value* OR (MH "Decision Making, Shared") OR shared decision making OR (MH "Decision Making) OR (MH "Decision Support Systems, Clinical") OR decision support) |
| 14. Heart Failure/<br>15. heart failure.ti,ab.<br>16. Ventricular Dysfunction, Left/<br>**17. or/14-16** | 14. Heart Failure/<br>15. heart failure.ti,ab.<br>16. Heart Ventricle Function/<br>17. ventricular dysfunction.ti,ab.<br>**18. or/14-17** | **2.** ((MH "Heart Failure+") OR (MH "Ventricular Dysfunction, Left+") OR heart failure) |
| 18. Drug Therapy/<br>19. medication.ti,ab.<br>20. drug.ti,ab.<br>21. pharmacotherapy.ti,ab.<br>22. Angiotensin-Converting Enzyme Inhibitors/<br>23. Angiotensin Receptor Antagonists/<br>24. sacubitril.ti,ab.<br>25. Adrenergic beta-Antagonists/<br>26. beta-blocker.ti,ab.<br>27. (carvedilol or bisoprolol or metoprolol).ti,ab.<br>28. Mineralocorticoid Receptor Antagonists/<br>29. Spironolactone/<br>30. Eplerenone/<br>31. Sodium-Glucose Transporter 2 Inhibitors/<br>32. (sglt2 or dapagliflozin or empagliflozin).ti,ab.<br>**33. or/18-32**<br>**34. 13 AND 17 AND 33** | 19. medication.ti,ab.<br>20. drug.ti,ab.<br>21. (drug therapy).ti,ab.<br>22. pharmacotherapy.ti,ab.<br>23. Dipeptidyl Carboxypeptidase Inhibitor/<br>24. (ace inhibitor).ti,ab.<br>25. Angiotensin Receptor Antagonists/<br>26. Sacubitril/<br>27. Sacubitril plus Valsartan/<br>28. Beta Adrenergic Receptor Blocking Agent/<br>29. beta-blocker.ti,ab.<br>30. (carvedilol or bisoprolol or metoprolol).ti,ab.<br>31. Mineralocorticoid Antagonist/<br>32. Spironolactone/<br>33. Eplerenone/<br>34. (spironolactone or eplerenone).ti,ab.<br>35. Sodium Glucose Cotransporter 2 Inhibitor/<br>36. Dapagliflozin/<br>37. Empagliflozin/<br>38. (sglt2 or dapagliflozin or empagliflozin).ti,ab.<br>**39. or/19-38**<br>**40. 13 AND 18 AND 39** | 3. ((MH "Drug Therapy+") OR pharmacotherapy OR (MH "Medication Treatment (Saba CCC)") OR (MH "Medication Management") OR medication)<br>4. **1 AND 2 AND 3** |

### Supplemental Appendix 2. Study data extraction instrument

| Data element | Description |
|---|---|
| Full study citation | |
| Reviewer identifier | - |
| Date extracted | - |
| Publication status | Whether obtained via database of published literature or grey literature |
| Objectives | Primary and secondary objectives/aims of the study |
| Years conducted | Timeframe of study conduct |
| Countries | Country(ies) where study was conducted |
| Methodology | e.g. Cross-sectional study, cohort, randomized controlled trial |
| Framework | Did the study use a decisional framework? If so, specify: _ |
| Sample size | n analyzed |
| Participants: Patients | Y/N |
| Definition/eligibility | |
| Recruitment strategy | |
| Sociodemographic factors (specific factors to extract, at minimum: Age; sex and gender; race or ethnicity; geography; education level; individual or household income; marital status; health or numerical literacy) | Assessed? Y/N<br>Specify: |
| Participants: Clinicians | |
| Type | e.g. Cardiologists, general practitioners |
| Recruitment strategy | |
| Decisional needs | Assessed Y/N |
| Difficult decision type/timing | Y/N, Specify details: |
| Unreceptive decisional stage | Y/N, Specify details: |
| Decisional conflict (uncertainty) | Y/N, Specify details: |
| Inadequate knowledge | Y/N, Specify details: |
| Unrealistic expectations | Y/N, Specify details: |
| Unclear values (option features) | Y/N, Specify details: |
| Inadequate support & resources (to make/implement the decision) | Y/N, Specify details: |
| Information overload | Y/N, Specify details: |
| Inadequate perceptions (others' views/practices) | Y/N, Specify details: |
| Difficult decisional roles | Y/N, Specify details: |
| Inadequate experience, self-efficacy, motivation, skills | Y/N, Specify details: |
| Inadequate emotional support, advice, instrumental help | Y/N, Specify details: |
| Inadequate financial assistance, health/social services | Y/N, Specify details: |
| Personal/clinical needs | Y/N, Specify details: |
| Others: | Y/N, Specify details: |
| Treatment preferences | Assessed? Y/N |
| HFrEF medications | General / specific class / specific drug |
| How? | Specify: |
| Attributes included | List (with rating/ranking/score): |
| Limitations: | |
| Notable author comments: | |
